# Development and validation of a deep-learning model to predict 10-year ASCVD risk from retinal images using the UK Biobank and EyePACS 10K datasets

**DOI:** 10.1101/2023.09.20.23295870

**Authors:** Ehsan Vaghefi, David Squirrell, Song Yang, Songyang An, Li Xie, Mary K. Durbin, Huiyuan Hou, John Marshall, Jacqueline Shreibati, Michael V McConnell, Matthew Budoff

## Abstract

**Background:** Atherosclerotic Cardiovascular Disease (ASCVD) is a leading cause of death globally, and early detection of high-risk individuals is essential for initiating timely interventions. The authors aimed to develop and validate a deep learning (DL) model to predict an individual’s elevated 10-year ASCVD risk score based on retinal images and limited demographic data.

**Methods:** The study used 89,894 retinal fundus images from 44,176 UK Biobank participants (96% non-Hispanic White, 5% diabetic) to train and test the DL model. The DL model was developed using retinal images plus age, race/ethnicity, and sex at birth to predict an individual’s 10-year ASCVD risk score using the Pooled Cohort Equation (PCE) as the ground truth. This model was then tested on the US EyePACS 10K dataset (5.8% Non-Hispanic white 99.9% diabetic), composed of 18,900 images from 8,969 diabetic individuals. Elevated ASCVD risk was defined as a PCE score of ≥7.5%.

**Results:** In the UK Biobank internal validation dataset, the DL model achieved area under the receiver operating characteristic curve (AUROC) of 0.89, sensitivity 84%, and specificity 90%, for detecting individuals with elevated ASCVD risk scores. In the EyePACS 10K and with the addition of a regression-derived diabetes modifier, it achieved sensitivity 94%, specificity 72%, mean error −0.2%, and mean absolute error 3.1%.

**Conclusion:** This study demonstrates that DL models using retinal images can provide an additional approach to estimating ASCVD risk, and the value of applying DL models to different external datasets and opportunities about ASCVD risk assessment in patients living with diabetes.

## Introduction

Atherosclerotic Cardiovascular Disease (ASCVD) is the most common cause of hospitalization and premature death in the US ^1^. The risk of an individual experiencing an ASCVD event includes both non-modifiable variables (age, sex, and race/ethnicity), and modifiable variables such as diabetes ^2^, hypertension ^3^, dyslipidemia ^4^, and smoking ^5^. Across a population, the risk of experiencing an ASCVD event varies greatly. Risk-based equations have therefore been developed to identify those who are at greatest risk of ASCVD so that preventive treatments can be initiated appropriate to the individual’s risk ^6^. The landmark Framingham Heart Study was the first to demonstrate that multivariable equations could identify an individual’s ASCVD risk with far greater accuracy than the existing metrics based solely on blood pressure and cholesterol ^7^. Since the Framingham-based equations were first published, other equations have been developed to serve different and more diverse populations with refined accuracy ^8–16^.

The retina is unique in being the only part of the human vasculature where the microvascular system is visible at micron-level resolution by non-invasive means. The automated detection of components within retinal images to predict ASCVD risk has been utilized with moderate degrees of success previously ^17–19^, but in recent years there has been an exponential increase in the number of studies that have used artificial intelligence (AI), and deep learning (DL) in particular, to extract data from retinal images ^20–22^. There is now growing interest in using the retinal image data generated by DL models to augment the traditional means of estimating ASCVD risk ^23^. In this study we used retinal photographs and limited demographic data from the UK Biobank to develop and validate a DL model designed to predict an individual’s elevated 10-year ASCVD risk, based on the US-derived Pooled Cohort Equation (PCE) ^24^.

The primary aim of this study was to develop and test a DL model to predict an individual’s 10-year ASCVD risk based on their retinal photographs plus age, race/ethnicity, and sex at birth and then further validate the findings in an external database. This model was built upon our previous work on detecting and grading retinopathy, maculopathy, macular degeneration, and effects of smoking in retinal images ^25–28^. While the Framingham risk score is recommended to perform cardiovascular risk assessment in some countries ^29^, the 2018 American Heart Association (AHA) Cholesterol Clinical Practice Guidelines recommend using the US-derived PCE to estimate the 10-year risk for hard ASCVD events (coronary heart disease death, nonfatal myocardial infarction, fatal or nonfatal stroke) ^24^. Our DL model was trained on and validated against the 10-year ASCVD risk as calculated by the PCE from individuals in the UK Biobank dataset (Level 3 in Figure 1, Appendix 2). Further external validation was provided by testing on a US-based dataset, EyePACS 10K, with substantially greater racial diversity and predominantly from patients living with diabetes.

**Figure 1:**
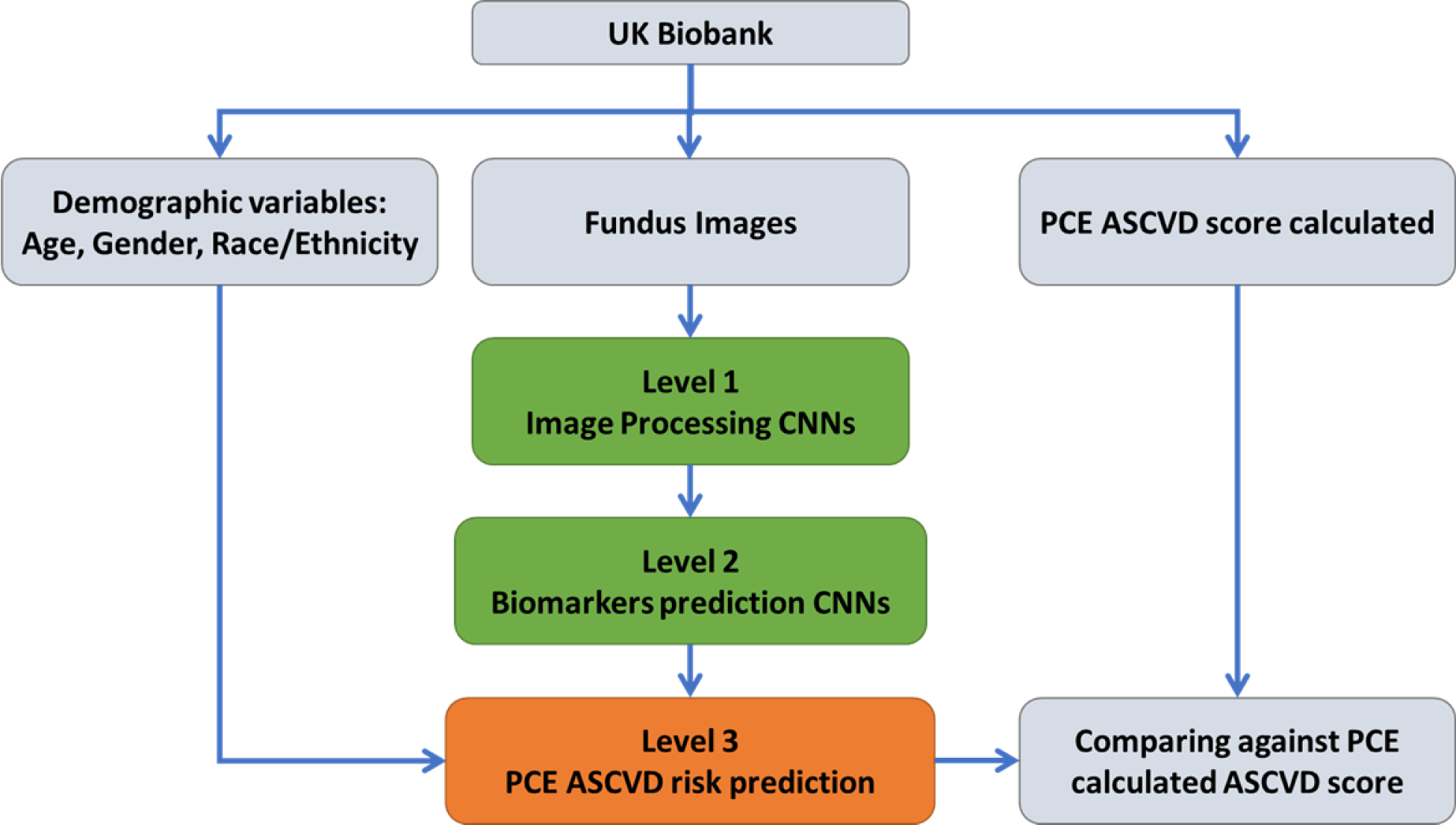
The process of training and validation of the DL prediction model using the UK Biobank dataset

## Methods

### Datasets

The composition of datasets used in this study is shown in Table 1. The UK Biobank was used for training and internal validation. The validation subset represented 20% of the data, selected randomly prior to development. The data from the UK Biobank can be accessed via a direct request to the UK Biobank; and was obtained using approved data management and data transfer protocols. 89,894 fundus images from 44,176 unique participants from the UK Biobank were used in this study. Participants in the UK Biobank were recruited from a UK general population with only approximately 5% of the UK Biobank population self-identified as having diabetes “diagnosed by doctor”.

**Table 1:** The demographic and risk factor makeup of the UK Biobank derived training and internal test datasets and the EyePACS 10K external validation dataset used in this study.

|  | UK Biobank:<br>Training<br>N = 35,570 |  | UK Biobank: Test<br>N=8,606 |  | EyePACS 10K<br>N=8,969 |  |
| --- | --- | --- | --- | --- | --- | --- |
|  | Mean | Std | Mean | Std | Mean | Std |
| Age (years) | 56 | 8.3 | 57 | 8.3 | 56 | 10 |
| Systolic Blood Pressure (mmHg) | 134 | 18.1 | 134* | 17 | 131** | 12 |
| Diastolic Blood Pressure (mmHg) | 81.4 | 10.1 | 81** | 9.8 | 71** | 8.9 |
| HbA1c (%) | 5.4% | 0.6% | 5.4%** | 0.5% | 8.1%* | 1.7% |
| Total cholesterol(mg/dL) | 220 | 44.1 | 219** | 43.2 | 179** | 43.7 |
| HDL cholesterol(mg/dL) | 58 | 15.1 | 58** | 15.1 | 47** | 12.3 |
| BMI | 27.2 | 4.7 | 27.1 | 4.6 | Not known | Not Known |
| Sex at birth | Male | Female | Male | Female | Male | Female |
|  | 16,313<br>(46%) | 19,257<br>(54%) | 3,991<br>(46%) | 4,615<br>(54%) | 3,923 (46%) | 5,046<br>(54%) |
| Current Smoker | TRUE | FALSE | TRUE | FALSE | TRUE | FALSE |
|  | 4,526<br>(13%) | 29,535<br>(87%) | 1,100<br>(13%)† | 7,122<br>(87%) | 507 (6%)† | 8,414<br>(94%) |
| Diabetes (%) | 5.02% |  | 4.78% |  | 99.95% |  |
| Race/Ethnicity | Non-Hispanic White | 93.23% | Non-Hispanic White | 93.04% | Non-Hispanic White | 5.8% |
|  | Asian without Chinese | 2.07% | Asian without Chinese | 2.13% | Asian | 6.5% |
|  | Chinese | 0.36% | Chinese | 0.27% | Asian sub- continent | 0.4% |
|  | Black | 2.14% | Black | 1.93% | Black | 6.8% |
|  | Hispanic | N.A. | Hispanic | N.A. | Hispanic | 65.7% |
|  | Mixed | 0.71% | Mixed | 0.80% | Mixed | 0.05% |
|  | Other | 1.16% | Other | 1.16% | Other<br>(including<br>Native<br>Americans) | 1.8% |
|  | Prefer not<br>to answer | 0.33% | Prefer not<br>to answer | 0.35% | Prefer not to<br>answer | 0.05% |
|  | Do not<br>know | 0.04% | Do not<br>know | 0.03% | Do not know | 12.9% |

As described below, a dataset from the US-based EyePACS study, with multiple differences from UK Biobank, was used for external validation. The dataset used for this analysis (EyePACS 10K) consisted of a subset of 9,947 individuals who had sufficient clinical data to calculate a traditional PCE risk score. Of these, 978 were excluded as they had established ASCVD prior to the date of retinal imaging. The external validation dataset thus comprised 18,900 images from 8,969 individuals.

The mean age of individuals in the EyePACS 10K dataset was 56±10 years (compared to 57±8.3 years in the UK Biobank). They predominantly self-identified as Hispanic whilst the UK Biobank population was predominantly non-Hispanic White. As the EyePACS 10K population consisted almost exclusively of people living with diabetes presenting for diabetic retinopathy screening, the mean Hemoglobin A1c (HbA1c) level of this population was high (8.1±1.7%). Additional demographics from both datasets are summarized in Table 1.

Significance test was performed between UK Biobank training and test, as well as Biobank training and EyePACS 10K external validation (* P<0.01 z test. **P<0.001 z test. † P<0.01 Chi square). The UK Biobank did not include Hispanic ethnicity as an option and the White participants were predominantly of British and Irish origin. EyePACS 10K included choices of Hispanic, Black, or White, so separating Hispanic Black participants from Hispanic White participants was not possible.

#### Inclusion/ Exclusion Criteria

Individuals in both datasets who had established ASCVD (Appendix 1) prior to the acquisition of the retinal images were excluded. A previously trained DL-based image quality model was used to screen all retinal images for acceptable image quality ^27^. Only the earliest acceptable images and the individual’s accompanying biometrics obtained on that same visit were used from the UK Biobank data. For the EyePACS 10K dataset, the earliest acceptable images which had accompanying biodata, taken no earlier than one year before image acquisition, were used. A detailed description of the DL model’s development is provided in Appendix 2.

### Model assessment

The performance of the DL model was assessed on both the 20% of UK Biobank that had been set aside for internal validation (8,606 individuals), and on all eligible patients in the EyePACS 10K external validation dataset (8,969 individuals).

#### Receiver operator curves (ROC), sensitivity, and specificity metrics

To assess the DL model’s ability to predict an elevated ASCVD score we compared the score it issued with that generated by the PCE equation. The DL model’s performance was based on a PCE score of ≥ 7.5% being the ground truth for elevated risk. The receiver operator characteristic curve (ROC), the precision recall curve, and the overall performance of the DL model on the UK Biobank and EyePACS 10K datasets were determined. The sensitivity and specificity of the model to detect individuals with an elevated ASCVD risk in the UK Biobank and EyePACS 10K datasets were also calculated. To investigate the performance of the model in different demographic groups in the UK Biobank, the performance of the model across sex, age, and race/ethnicity were also calculated. To investigate the impact the retinal image had on the DL model’s performance, and to ascertain whether the retinal data contributed positively to the output, we re-ran the DL model withholding the retinal image input data and recalculated the AUROC, sensitivity, and specificity.

#### Comparison of the performance of the DL model and the PCE equation to predict 10-year ASCVD risk and events

The performance of the DL model vs. PCE was also assessed by way of a data binning technique ^30^. Patients in the UK Biobank and EyePACS 10K datasets were first arranged in ascending order by their DL model predicted scores and then by their PCE-calculated 10-year ASCVD risk scores. Both datasets were then divided into 20 bins with equal population numbers by an ascending order of their ASCVD risk scores, i.e., from the lowest 5%, every 5% to the top 5%. The mean 10-year ASCVD risk scores generated by the DL model and the PCE equation were then plotted for each bin in both the UK Biobank and EyePACS 10K datasets. The magnitude of the deviation between the two sets of results generated for each dataset was then assessed by measuring the mean error and the mean absolute error per case.

For the UK Biobank the exercise was then repeated using actual observed ASCVD events, categorized by risk as determined by the DL model and the PCE equation. This exercise was not possible for the EyePACS 10K dataset as there were no future ASCVD events recorded after the retinal images were acquired. In accordance with ACC/AHA Work Group guidelines, an ASCVD event for the purposes of outcomes was defined as the first non-fatal acute myocardial infarction, fatal or non-fatal stroke, or fatal coronary heart artery disease ^11^. The ICD codes used to define a hard ASCVD event are provided in Appendix 1.

Finally, to assess the probabilistic performance of the two methods for predicting 10-year ASCVD risk, the Brier score loss was calculated for both the DL model and the PCE equation.

## Results

The ability of the DL model to identify elevated-risk individuals (PCE-generated ASCVD score ≥7.5%), compared to the PCE equation, for both the UK Biobank and EyePACS 10K datasets are shown in Table 2. In UK Biobank, the DL model achieved AUROC: 0.89, a sensitivity of 83%, and a specificity of 90%. Withholding the retinal image data from the DL model resulted in lower performance, with AUROC 0.84, sensitivity 70% specificity 88%.

**Table 2:** Confusion matrices comparing the DL model-predicted ASCVD score v PCE-calculated ASCVD scores. The numbers in each cell are numbers of people.

|  | UK BioBank |  | EyePACS 10K |  | EyePACS 10K + diabetes modifier |  |
| --- | --- | --- | --- | --- | --- | --- |
|  | PCE-generated ASCVD score <7.5% | PCE-generated ASCVD score ≥7.5% | PCE-generated ASCVD score <7.5% | PCE-generated ASCVD score ≥7.5% | PCE-generated ASCVD score <7.5% | PCE-generated ASCVD score ≥7.5% |
| DL model generated ASCVD score <7.5% | 5923 (69%) | 331 (4%) | 4126 (46%) | 2238 (25%) | 3015 (34%) | 279 (3%) |
| DL model generated ASCVD score ≥7.5% | 642 (7%) | 1710 (20%) | 217 (2%) | 2388 (27%) | 1328 (15%) | 4347 (48%) |

In EyePACS10K, the DL model achieved a similar AUROC: 0.90, with a lower sensitivity of 52% and a higher specificity of 95%. The ROC and the accompanying precision-recall curve plots are shown in Figure 2.

**Figure 2:**
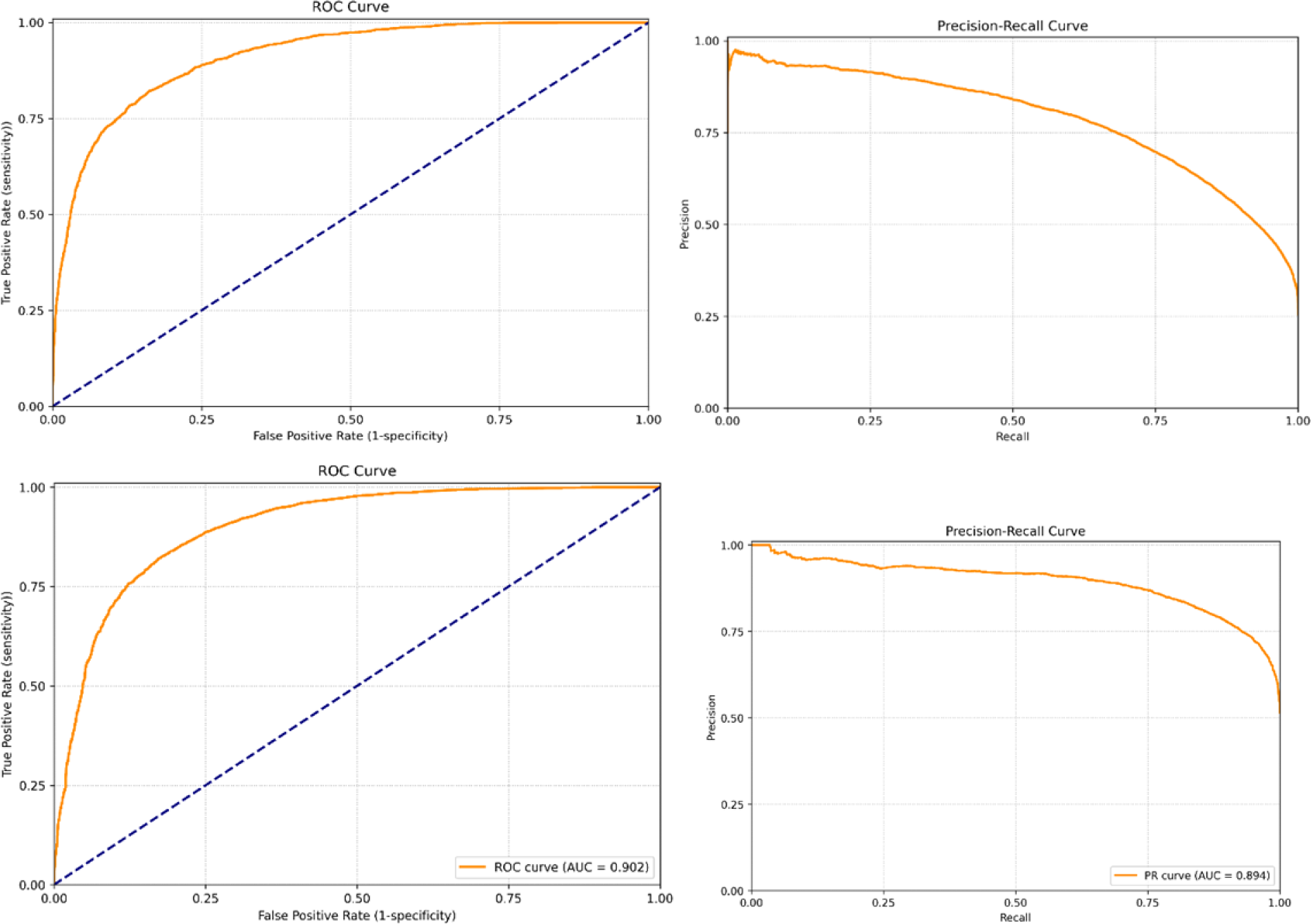
(left) the receiver operating characteristic (ROC) curve, (right) the precision-recall curve, for UK Biobank (top) and for EyePACS 10K (bottom row)

In post-hoc analysis, applying a regression-based diabetes modifier to the DL model output, the sensitivity and specificity to detect individuals with elevated risk against PCE in the EyePACS10K dataset were 94% and 72% respectively.

The AUROC values were then calculated for each demographic segment of both the UK Biobank and EyePACS 10K datasets [Tables 3a, 3b and 3c]. The DL model performed well across both sexes and all races/ethnicities, but its performance in younger individuals (≤ 60yrs) in UK Biobank was lower than older individuals (AUROC 0.72 v 0.84).

**Table 3a:** Performance of DL model (AUROC) across different race/ethnicities and demographics in the UK Biobank test dataset.

| Demographic segment | | AUROC | Group size | N where PCE $\geq 7.5\%$ |
| --- | --- | --- | --- | --- |
| Sex at birth | Female | 0.88 | 4615 | 217 |
|  | Male | 0.89 | 3991 | 1824 |
| Race/Ethnicity* | White | 0.87 | 8023 | 1945 |
|  | Asian (including Chinese) | 0.83 | 226 | 45 |
|  | Black | 0.89 | 171 | 29 |
|  | Others | 0.93 | 208 | 31 |
| Age bracket | Age $\leq 60$ | 0.72 | 5112 | 417 |
| | Age $> 60$ | 0.84 | 3494 | 1624 |

\*Participants self-identified as belonging to a particular race/ethnicity group (Appendix 3)

**Table 3b:** Performance of DL model (AUROC score) across different race and demographics in EyePACS 10K dataset.

| Demographic Segment | | AUROC | Segment Size | N where PCE ≥ 7.5% |
| --- | --- | --- | --- | --- |
| Sex at birth | Female | 0.89 | 5352 | 1975 |
|  | Male | 0.86 | 3612 | 2648 |
| Race/ethnicity | Hispanic | 0.91 | 5889 | 2814 |
|  | Black or African American | 0.85 | 609 | 426 |
|  | Non-Hispanic White | 0.91 | 516 | 323 |
|  | Asian | 0.9 | 586 | 361 |
|  | not specified | 0.92 | 1158 | 575 |
|  | Indian subcontinent origin | 0.95 | 37 | 18 |
|  | Others | 0.89 | 157 | 100 |
| Age bracket | Age ≤ 60 | 0.88 | 5572 | 1751 |
| | Age $>$ 60 | 0.84 | 3392 | 2872 |

**Table 3c:** Performance of DL model (AUROC score) across different ethnicities/races and demographics in EyePACS 10K dataset when diabetes modifier is applied.

| Demographic Segment | | AUROC | Segment Size | N where PCE ≥ 7.5% |
| --- | --- | --- | --- | --- |
| Sex at birth | Female | 0.92 | 5352 | 1975 |
|  | Male | 0.91 | 3612 | 2648 |
| Race/ethnicity | Hispanic | 0.94 | 5889 | 2814 |
|  | Black or African American | 0.87 | 609 | 426 |
|  | Non-Hispanic White | 0.92 | 516 | 323 |
|  | Asian | 0.92 | 586 | 361 |
|  | not specified | 0.94 | 1158 | 575 |
|  | Indian subcontinent origin | 0.97 | 37 | 18 |
|  | Others | 0.92 | 157 | 100 |
| Age bracket | Age ≤ 60 | 0.9 | 5572 | 1751 |
| | Age $>$ 60 | 0.86 | 3392 | 2872 |

\* Race/ethnicity was self-reported by study participants (Appendix 3)

### Relationship between ASCVD risk scores generated and observed ASCVD event rates using the traditional PCE score, and the DL model predicted score

#### 1. UK Biobank

The predicted 10-year ASCVD risk scores generated by the PCE equation and the DL model, plotted for each bin in the UK Biobank, are shown in Figure 3a. The predicted 10-year ASCVD risk scores rose equally and proportionally for all risk categories in both cohorts. Across the UK Biobank, the predicted 10-year ASCVD risk scores from PCE and the DL model were very similar (mean error 0.3%, mean absolute error 2.4%). The actual ASCVD event rates observed in both the PCE and DL model, when categorized by ascending order of predicted risk, are also shown in Figure 3a. The actual ASCVD event rate rose steadily from the non-elevated to elevated-risk categories in both the PCE and DL models across all individuals in the UK Biobank. The magnitude of the actual ASCVD event rates were again very similar to the predicted 10-year ASCVD risk score produced by the PCE and DL models for all risk categories. The actual ASCVD event rates observed when the results generated by the PCE and DL models were subdivided into the binary classification (predicted risk score <7.5 % or ≥7.5%) are shown in Table 4. The ASCVD event rate observed in those individuals from the UK Biobank allocated a “non-elevated” score by the traditional PCE method was the same as those allocated a “non-elevated” score by the DL prediction model (2.2% v 2.0%). The same finding was observed in the “elevated” risk groups; (ASCVD event rate: traditional PCE 7.5%, DL model 7.5%). Analysis with the Point Biserial Correlation Coefficient (PBCC) revealed that the 10-year ASCVD risk score produced by the PCE and the DL model were both significantly correlated with actual ASCVD events. Calculated PCE vs. ASCVD events: 0.144 (P<0.01); DL model predicted risk score vs. ASCVD events: 0.152 (P< 0.01). The accuracy of the PCE equation to correctly predict an ASCVD event was identical to that of the DL model, with the Brier score loss for both 0.067.

**Figure 3a:**
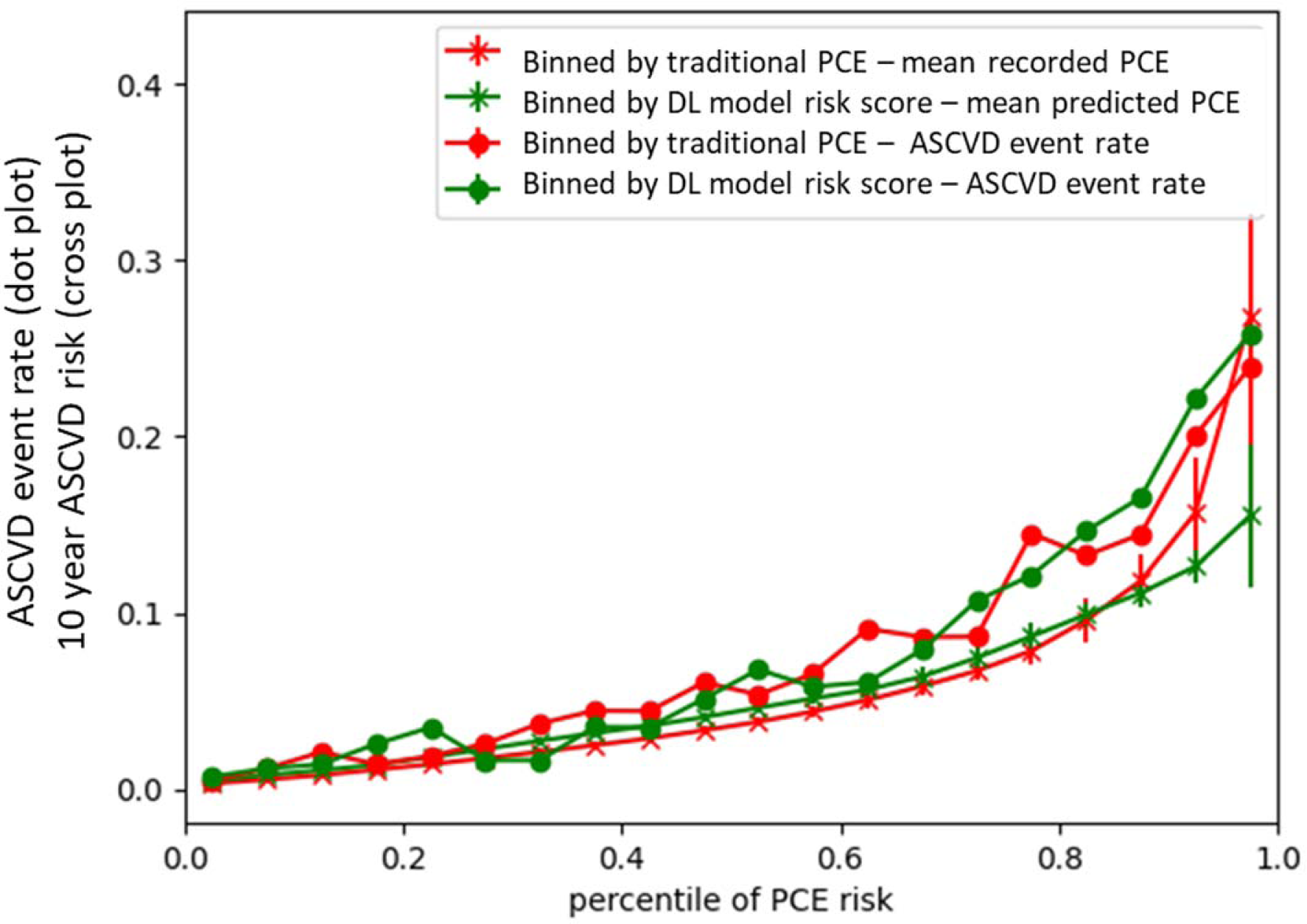
10-year ASCVD risk scores (crosses) and ASCVD event rates (dots) when categorized according to the traditional PCE-calculated risk score (red) or DL model predicted risk score (green) in the UK Biobank.

**Table 4.** UK Biobank ASCVD event rates in those with and without elevated ASCVD risk scores by PCE-based ASCVD risk score compared to DL model-predicted ASCVD risk score. The numbers in each cell are #people (#ASCVD events)/% events/cases.

| 10-year ASCVD risk score:<br>PCE | Individuals<br>(ASCVD events) | 10-year ASCVD risk score:<br>DL model | Individuals<br>(ASCVD events) |
| --- | --- | --- | --- |
| <7.5% | 6565 (146)<br>2.2% | <7.5% | 6254 (123)<br>2.0% |
| ≥7.5% | 2041 (153)<br>7.5% | ≥7.5% | 2352 (176)<br>7.5% |

#### 2. EyePACS 10K dataset

The ASCVD risk scores generated by the PCE equation and the DL model, plotted for each bin in the EyePACS 10K dataset, are shown in Figure 3b. For all risk categories the predicted 10-year ASCVD risk score, as measured by the PCE equation, was substantially higher than that produced by the DL model (mean error 3.6%, mean absolute error 4.4%). Comparison between Figures 3a and 3b reveals that the performance of the DL model was very consistent across both the UK Biobank and EyePACS 10K. However, the PCE equation predicted consistently higher scores across all risk profiles in the EyePACS dataset, unlike in UK Biobank.

**Figure 3b:**
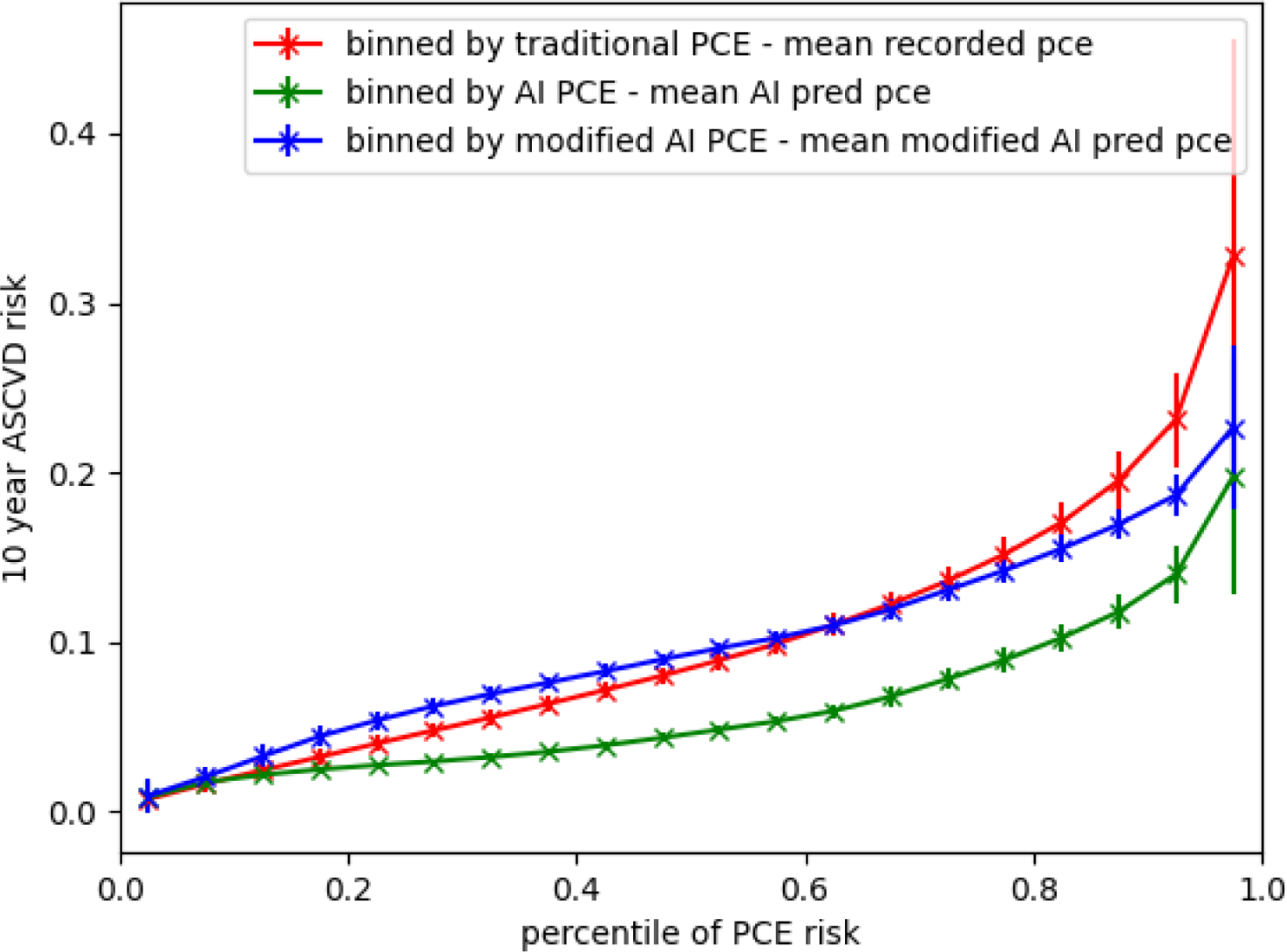
10-year ASCVD risk when categorized according to the traditional PCE-calculated risk score (red) or DL model-predicted risk score (green) or DL model-predicted score after application of the diabetes modifier (blue) in the EyePACS 10K dataset.

As the ROC curves indicated that the performance of the DL model was very similar in both the internal (AUROC 0.89) and external (AUROC 0.90) validation datasets, but with differing sensitivities and specificities, we hypothesized that there was a systematic difference between the 10-year ASCVD risk score produced by the PCE and our DL model when it was applied to people living with diabetes. If so it should be possible to produce a correction factor; a “diabetes modifier,” which would translate the 10-year ASCVD risk score produced by our DL model to that produced by the PCE, across all risk profiles ^31^. The regression equation that solved this systematic difference was: DM-modified ASCVD risk score = DL model ASCVD risk score + max (diabetes_adjustment, 0), where diabetes_adjustment= −10.4156+(0.3560*age)+(−4.5422*gender)+(−0.5410*pce_pred). For sex, Male = 0, Female = 1. pce_pred range [0-100]. The ASCVD risk score generated by DL model with the diabetes modifier applied, plotted for each bin in the EyePACS 10K dataset, is shown in Figure 3b. The results produced by the DL model with the diabetes modifier applied were now very similar to PCE (mean error −0.2%, mean absolute error 3.1%). The sensitivity and specificity of the new diabetes-modified DL Model to detect individuals with elevated risk against PCE in the EyePACS10K dataset were 94% and 72% respectively.

## Discussion

In this study we developed and validated a novel DL model to calculate a 10-year ASCVD risk score using over 90,000 retinal images from the UK Biobank dataset and then externally validated it on over 18,000 retinal images from the EyePACS 10K dataset. Our DL model reliably detected those individuals with elevated ASCVD risk scores (≥7.5%) using only the retinal image and the individuals’ age, race/ethnicity, and sex with an AUROC of 0.89, a sensitivity and specificity of 84% and 90% respectively in UK Biobank and an AUROC of 0.90, a sensitivity and specificity of 52% and 95% respectively in EyePACS 10K.

Participants in the UK Biobank were recruited from a UK general population and were mainly non-Hispanic White. Individuals in the US-based EyePACS 10K dataset predominantly identified as Hispanic and the majority were people living with diabetes presenting for diabetic retinopathy screening. This makes direct comparison with respect to ASCVD risk predictions between the two datasets challenging. As such it is important to also examine the performance of our DL model and the PCE in the context of actual ASCVD events. The actual observed ASCVD event rate in UK Biobank was very similar between the DL model and PCE; both had ∼2% ASCVD event rate observed in those allocated “non-elevated” risk, and 7.5% in those allocated to the “elevated”-risk group. There was also a significant correlation between the risk scores produced by both models and actual ASCVD events (PCE-based risk score v ASCVD events 0.144, P<0.01; DL model-predicted risk score v ASCVD events 0.152, P< 0.01), and the probabilistic accuracy of the DL model to correctly predict an ASCVD event was identical to that of the PCE (Brier score loss 0.067).

There was a clear difference between the PCE and the DL model-predicted 10-year ASCVD risk scores in the EyePACS 10K dataset. When assessed by the PCE, 48% of individuals in the EyePACS 10K dataset were deemed to be “non-elevated” and 52% were deemed “elevated” risk. The DL model apportioned the risk differently: 70% “non-elevated”; 30% “elevated” risk. As the PCE score was regarded as the ground truth in the confusion matrix, this difference in how the two models apportioned risk explains the apparently low sensitivity of the DL model. Data binning is a method to group continuous values into a smaller number of “bins’’ and is commonly used by data scientists to reduce the effects of observation errors [30]. This approach was used to further investigate the difference in how the PCE and our DL model apportioned risk in both the UK Biobank and EyePACS 10K datasets, plotting the 10-year ASCVD risk when categorized according to the traditional PCE-calculated risk score or DL model predicted risk score. Unlike what was observed in the UK Biobank, when the 10-year ASCVD risk scores generated by the PCE and the DL model from the EyePACS 10K dataset were grouped into bins of ascending risk scores, the magnitude and distribution of the predicted 10-year ASCVD risk scores generated by the PCE and the DL model were quite different; mean error 3.6%, mean absolute error as measured by each index case was 4.4% [Figures 3b]. As the performance of the DL model assessed by the AUROC was very similar in both the UK Biobank (0.89) and EyePACS 10K (0.90) datasets, and the principle difference in the two datasets was low vs. high rate of diabetes, we hypothesized that it should be possible to regress a correction factor; a “diabetes modifier,” to transform the ASCVD risk score produced by our DL model to that produced by the PCE. Applied to the EyePACS 10K dataset, this post-hoc analysis showed that 10-year ASCVD risk scores generated by the DL model were substantially better aligned to the PCE after the diabetes modifier was applied: mean error −0.2% and mean absolute error 3.1% [Figure 3b]. The subsequent sensitivity and specificity of the “diabetes-modified” DL model to detect individuals with “elevated” risk in the EyePACS10K dataset were also improved at 94% and 72% respectively. Although further validation in other datasets is required, these findings indicate that by applying an appropriate “diabetes modifier”, our DL model can more accurately match the 10-year ASCVD risk produced by the PCE when used in people living with diabetes.

Regardless of diabetes type and status, the PCE equation currently treats all people living with diabetes as a uniform group and it effectively adds a modifier to the regression equation which serves to elevate their risk score compared to non-diabetics. Recently it has been suggested that the traditional regression-based equations, like the PCE, may overestimate ASCVD risk in many people living with diabetes ^32,33^. An individual living with longstanding poorly controlled type 1 diabetes for example may have a very different vascular risk profile to an older person who has just been diagnosed with mild type 2 diabetes, and yet the PCE does not discriminate between the two; both “have diabetes”. Tools that better discriminate ASCVD risk in people living with diabetes are therefore required. Recently it has been demonstrated that retinopathy could be used to further refine ASCVD risk in people living with diabetes. Modjtahedi et al demonstrated that not only is diabetic retinopathy associated with future risk of ASCVD events and death, but the magnitude of this risk correlates with the level of retinopathy ^34^. The authors hypothesize that retinopathy could be representative of wider macrovascular disease and conclude that future calculators, designed to stratify ASCVD risk in people living with diabetes, should incorporate detailed retinopathy input information to more accurately model macrovascular risk. Presently this retinopathy data can only be made available to developers and users of ASCVD risk calculators if the subject has a recent retinal image which has been appropriately graded. DL models like ours, designed to predict ASCVD risk based on retinal images and trained on datasets which contain detailed retinopathy results, could provide the ideal opportunity to capture this retinopathy data and incorporate these findings into ASCVD risk prediction. Further studies, designed to corroborate retinopathy findings and ASCVD risk scores with actual ASCVD events in individuals living with diabetes are therefore required. Until newer risk calculators that incorporate retinopathy data into the risk prediction tool are made available, we have demonstrated that it is possible to train a DL model on a general population and overlay a diabetes modifier which then enables it to more accurately and consistently match the ASCVD risk scores issued by the PCE to people with diabetes.

The application of DL algorithms to predict ASCVD risk-related outcomes from retinal images has been comprehensively reviewed by Hu et al. ^35^. This review revealed that to date a heterogeneous array of models have been developed using a variety of different inputs: retinal images only, retinal images + various biodata, and reporting against different cardiac-related outcomes. Direct comparison between different models is therefore challenging, but one common theme to emerge was that few of these models have been validated on external datasets. Even fewer have utilized an external dataset derived from individuals with a very different racial/ethnic background. In the current study we trained and tested a DL model to predict the 10-year risk of ASCVD events in the UK Biobank and then further validated the results in an external database which had a very different racial/ethnic make up. To date only two other groups have reported the results of a DL model trained and then externally validated to predict the ASCVD 10-year risk from retinal photographs ^22,36,37^. Our results support the accumulating evidence that indicates DL algorithms can use retinal images to accurately predict ASCVD risk and they compare favorably with the one other DL model that has been validated on the UK Biobank; which had sensitivity, specificity of 83%, 88%, respectively ^22,36,37^.

### Strengths and Limitations

The primary strengths of this study include: we have trained and validated a DL model designed to predict an individual’s ASCVD 10-year risk based on nothing more than a retinal photograph and limited demographic data; the DL model was not only able to reliably match the PCE scores, but also showed similar prediction to PCE of actual ASCVD events; and the DL model was externally validated on a very different dataset. The EyePACS 10K dataset, which although very similar in age, differed substantially in diabetes prevalence, ethnic/racial makeup, and geography (US vs. UK). Testing of the model across race/ethnicity and demographics within the test dataset revealed that the DL model performed consistently well across most groups. However, its ability to predict ASCVD risk was less accurate for individuals of “other races” and those individuals aged less than 60. The former probably reflects the fact that there are a relatively low number of individuals of other races represented in the UK Biobank dataset. The latter may reflect an age bias in that ASCVD risk increases with, and is strongly correlated, to age. Thus, in common with all DL models, one fundamental weakness of the current study is that our model may have limited generalizability to other datasets whose racial and demographic makeup differ to those seen in the UK Biobank and our results demonstrate the importance of ensuring that DL models designed for health care settings should be trained and validated in the designated target population. Our DL model performed similarly (by AUROC) on the external EyePACS10K dataset as it did on the UK Biobank, and we investigated the application of a “diabetes modifier” to the output of our DL model to better match the sensitivity and specificity of the PCE when applied to individuals living with diabetes. However, as there were no future hard ASCVD events in the EyePACS 10K dataset, we could not validate the DL model on this key metric. Our results will therefore need to be replicated on other external datasets before it can be considered a clinically useful tool.

In the development of our DL model we relied on a systolic hypertension DL model as the sole measure of blood pressure. It is feasible that a model that combined both SBP and diastolic blood pressure may have yielded more accurate results than SBP alone. Finally, it is highly probable that the ground truth for the smoking DL model, namely self-reported smoking status, lacks robustness as it is widely accepted that individuals tend to underreport their smoking habit ^38,39^. As the PCE equation places a high weighting on an individual’s smoking status, any deficiencies in the smoking model could have a significant impact on the final predicted ASCVD risk.

## Conclusion

In conclusion, our results show that it is possible to train a DL model that can assess ASCVD risk as well as the traditional PCE method, using nothing more than a retinal photograph and limited demographic data. We have also shown that the application of a “diabetes modifier” to our DL model is a promising approach to matching the PCE in people living with diabetes. If these results can be replicated in other large population-based datasets, DL models like ours may offer the potential to significantly improve access to ASCVD risk detection strategies as the risk predictions these models produce do not require multiple clinical and laboratory assessments to generate an individual’s ASCVD risk score. As retinal photographs are routinely captured in optometry, ophthalmology, and some pharmacy clinics, it means these DL models can be deployed without significant additional investment, making this technology particularly relevant to lower-resource settings. As an additional ASCVD risk assessment screening tool, a DL prediction model has the potential to make initial ASCVD risk assessment more affordable and accessible to all, with the goal of improving patient outcomes through earlier detection and increasing the public awareness of ASCVD risk and prevention.

## Data Availability

Databases used in this study are obtainable from their original data guardian, UK Biobank and EyePACS.

## Appendix 1

**Definition of “clinical” ASCVD event used to identify events prior to the acquisition of the retinal image. This list defines exclusion criteria for the validation datasets.**

**Acute myocardial infarction** was defined by ICD-9-CM codes 410.* and ICD-10-CM codes I21.*, I22.*, I23.3, I24.0, I24.9, I25.9, or I51.3

**Angina:** was defined by ICD-9-CM codes 413.* and ICD-10-CM codes I20.*, I23.7, I25.7, I125.11x

**CABG/ PCI:** were defined based on ICD-9-CM codes 414.02, 414.03, 414.04, 414.05, V45.81, V45.82 and ICD-10-CM codes Z95.1, T82.21.21*, I25.7* (exclude I25.75), I25.810, I25.812, Z98.61, Z95.5

**Stroke events** were defined based on ICD-9-CM codes 430.*, 431.*, 432.*, 433.*1, 434.*1, or 436.0 and ICD-10-CM codes G45, G46.*, I63.*, I67.85, I69.30, I77.89, P91.0, or Z86.73.

**Definition of hard ASCVD event used to detect events subsequent to acquisition of the retinal image in the validation test datasets**

**Acute myocardial infarction** was defined by ICD-9-CM codes 410.* and ICD-10-CM codes I21.*, I22.*, I23.3, I24.0, I24.9, I25.9, or I51.3

**Stroke events** were defined based on ICD-9-CM codes 430.*, 431.*, 432.*, 433.*1, 434.*1, or 436.0 and ICD-10-CM codes G46.*, I63.*, I67.85, I69.30, I77.89, P91.0, or Z86.73.

**Fatal coronary artery disease was defined by the presence of** an ICD-9-CM code 411.*, 413.*, or 414.* or an ICD-10-CM code I20.*, I23.7, I24.*, I25.*, or T82.85 code followed by death within a year.”

## Appendix 2

### Model development

The DL model pipeline encompasses approximately 50 distinct DL models, classified into two primary categories: image-based models and non-image-based models. The former uses image data as input, while the latter relies on vectorized interpretations of participant information including their biometrics. This dichotomy results in a diversity of input data and target formats within the pipeline. For instance, the systolic blood pressure (SBP) model utilizes a retinal image as input, with the corresponding SBP value serving as the target. This model’s function is independent of additional factors, thereby enabling it to be trained on samples missing the HbA1c value, provided the SBP data are accessible. Another model of note is the PCE ASCVD prediction model. It uses outputs from preceding image-based models, with the calculated PCE results serving as the target. However, the PCE equation requires multiple variables, including SBP, HbA1c, total cholesterol, and high-density lipoprotein (HDL) cholesterol. Only participants with all of the above-mentioned components available were used in this study.

### Model ensemble

The DL prediction model ensemble used is demonstrated in Figure 2 and comprises 3 different levels:

**Figure 1:**
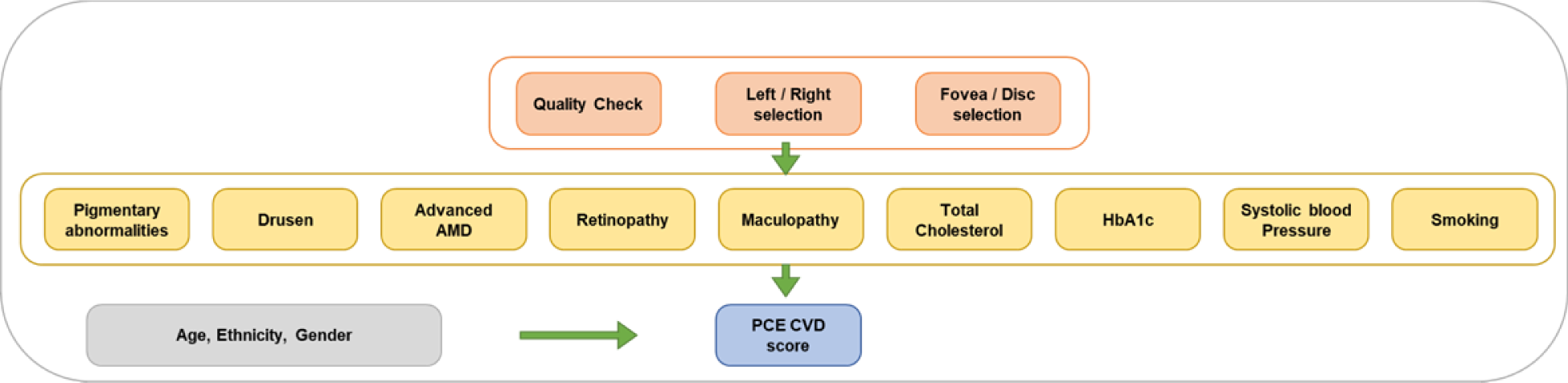
The structure of DL prediction model developed in this manuscript

#### Level 1

The first level includes an image quality check CNN (described above), a laterality (Left eye/Right eye) detector CNN, and an image location (fovea / non-fovea) detector CNN. The input of this layer is retinal images only. This process ensures that only foveal-centered images that are of sufficient quality are accepted into the model. Identifying the laterality of the image ensures that only a single fovea-centered image for each eye for each individual is used during the analysis. The final available dataset after requiring high- or medium-quality, fovea-centered images, one per eye, consisted of 89,894 images, representing 44,176 patients. This was divided into an 80/20 split for training and validation, respectively. The training dataset thus consisted of 76,321 images (representing 35,570 patients). Meanwhile, the test dataset comprised 19,080 images representing 8,606 patients who had valid biometrics. The demographics of the final dataset did not significantly differ from those that included images of low quality. The patient demographics of the test and training datasets were statistically compared with a two-sample Kolmogorov-Smirnov test to ensure that the demographic distribution of the training and test group were similar. There was no statistically meaningful difference between the two datasets, comparing the age, gender and race makeup of the two groups [Supplementary material]. The training of these models is explained elsewhere, and these models were not tuned or retrained for this study [26].

#### Level 2

The second level includes nine ensembles of AIs, each consisting of five CNNs (45 in total). The retinopathy, maculopathy, drusen, pigmentary abnormality, advanced AMD, and smoking CNNs were previously trained on other datasets [25-28]. The rest of the CNNs were trained using the unique UK Biobank labels in the fundus images:′ ′hba1c_result′, ′tchdl_result′, ′systolic_bp′, ′systolic_bp2′, ′smoking_status.′ Systolic_bp′ and ′systolic_bp2′ are two consecutive blood pressure measurements in the UK Biobank and in this study we used the mean of the two.

These CNNs follow modified versions of the Inception-Resnet-V2 or ResNet50 structures. Taking the single retinopathy CNN model as an example, the model has a deep structure, consisting of 164 layers, and uses a combination of inception and residual blocks. The inception blocks use a combination of convolutional layers with different filter sizes, while the residual blocks use skip connections to enable the model to learn from previous layers. We also employed batch normalization and bottleneck layers to improve training efficiency. Overall, the model architecture is designed to extract features at multiple scales and capture fine-grained details in images, making it well-suited to detect the level of retinopathy or other biomarkers. For each CNN in Level 2, the image plus biomarkers dataset was split for training, validation, and testing: 70%, 15%, 15% respectively. The excessive background of the fundus images was cropped, and the resulting image was resized to 800×800 pixels. A batch size of 8 was chosen to optimize GPU memory during training. Adam optimizer was adopted with a learning rate 1*10e-3 to update parameters towards the minimization of the loss. Dropout was enabled with a rate p = 0.2, and the model was trained for at least 100 EPOCHs. All codes related to this work were implemented using Python 3.7.

Additionally, we developed a complex jury system to arrive at the ultimate prediction for each biomarker. To elaborate, using the retinopathy model as an example, there exist six distinct levels of retinopathy (R0-R5). Five jury models were employed to assess each eye, resulting in 30 probability values per eye. These probabilities were merged and consolidated for both eyes, thereby yielding a final value for each patient.

#### Level 3

The third level is a Multi-Layer Perceptron (MLP), which uses the output of the second level CNNs, plus the patient’s chronological age, gender, and race to estimate their PCE ASCVD risk score. This PCE-derived ASCVD risk score is the ground-truth label, calculated from the relevant 9 fields in the UK Biobank dataset for each participant (age, sex, race, smoking status, blood pressure, diabetes, serum total cholesterol, HDL cholesterol, blood pressure lowering medication https://tools.acc.org/asASCVD-risk-estimator-plus/#!/calculate/estimate/). The architecture of the model comprises an input layer, followed by five dense layers that exhibit a gradual decrease in neuron counts, namely 1024, 512, 256, 128, and 32. These layers are interspersed with batch normalization and LeakyReLU activation functions with a leaky rate of 0.1. To address overfitting concerns, dropout layers with a rate of 0.3 were incorporated after the third, fourth, and fifth dense layers. The ultimate layer, encompassing a single neuron and a linear activation function, predicts the target value. For optimization purposes, an Adam optimizer is utilized with an exponentially decaying learning rate schedule, initialized at 3e-3 and decaying by a factor of 0.95 every 1000 steps. The Huber loss function was employed to guide the model parameters updating. To curb overfitting and ensure efficient training, early stopping was implemented.

## Appendix 3

The summary list of ethnicities for the UK Biobank and EyePACS 10k studies

| Summary List | UK Biobank List | EyePACS 10k List |
| --- | --- | --- |
| Non-Hispanic White | White (predominantly British/Irish) | Caucasian/White |
| Hispanic* | N.A. | Latin American/Hispanic |
| Black/African American* | Black | African/African American |
| South Asian | Asian (predominantly South Asian) | Indian subcontinent |
| East Asian | Chinese | Asian/Pacific Islander |
| Native American | N.A. | Native American/Alaskan Native |
| Multi-racial | Mixed | Multi-racial |
| Other | Other | Other |
| Declined/Do not know | Prefer no answer/do not know | Decline to state |

\* UKB did not capture Hispanic ethnicity. EP had reporting options of Hispanic, Black, or White, so it was not possible to report Hispanic Black and Hispanic White separately.

